# Longitudinal functional network connectivity changes across the clinical stages of *C9orf72* hexanucleotide repeat expansion carriers

**DOI:** 10.1101/2025.11.23.25340528

**Authors:** Liwen Zhang, Suvi Häkkinen, Youjin Jung, Maria Luisa Mandelli, Dana Leichter, Chiadi U. Onyike, Julio C. Rojas, Jolina Lombardi, Maria Luisa Gorno-Tempini, Jennifer S. Yokoyama, Virginia E. Sturm, Joel Kramer, Brad F. Boeve, Adam L. Boxer, Leah K. Forsberg, Hilary W. Heuer, Kejal Kantarci, Eliana Marisa Ramos, Howard J. Rosen, Bruce L. Miller, William W. Seeley, Taru M. Flagan, Suzee E. Lee, the ARTFL/LEEFTDS/ALLFTD Consortia

## Abstract

**INTRODUCTION:** Intrinsic functional connectivity network abnormalities in *C9orf72* hexanucleotide repeat expansion carriers emerge during the asymptomatic phase, yet longitudinal studies remain limited. We examined cross-sectional abnormalities and longitudinal connectivity changes across clinical stages.

**METHODS:** We analyzed task-free fMRI and structural MRI data in 36 asymptomatic (aSxC9), 17 prodromal (proC9), and 29 symptomatic (SxC9) carriers, and 107 healthy controls (HC). Functional networks previously found altered in *C9orf72*, including salience, sensorimotor, default mode, and medial pulvinar thalamic networks, were examined. Associations between longitudinal connectivity and gray matter decline with baseline neurofilament light chain (NfL) concentrations and symptom severity were assessed.

**RESULTS:** Despite lacking detectable gray matter decline, aSxC9 and SxC9 showed longitudinal connectivity changes within specific networks. In aSxC9, connectivity changes correlated with baseline NfL. In proC9 and SxC9, changes in connectivity and gray matter were associated with baseline NfL and symptom severity.

**DISCUSSION:** *C9orf72* expansion carriers demonstrate stage-specific network connectivity changes.

**Highlights:**

- Stage-specific longitudinal connectivity changes were detected in *C9orf72*.
- Compared to controls, each *C9orf72* cohort lacked detectable gray matter decline.
- For aSxC9, connectivity network changes correlated with baseline NfL.
- Connectivity/GM changes in proC9 and SxC9 correlate with NfL and symptom severity.

**Research in Context:** Systematic review: The authors conducted a systematic review using major databases (e.g., PubMed and Google Scholar). While cross-sectional studies have investigated functional connectivity patterns in *C9orf72* expansion carriers (*C9orf72*), longitudinal studies are scarce. Notably, no study has examined longitudinal functional connectivity changes during the prodromal stage, a critical transition stage from the asymptomatic to symptomatic phases, which was included in this study.

Interpretation: Leveraging a large longitudinal neuroimaging cohort of *C9orf72*, we comprehensively characterized stage-specific connectivity changes in asymptomatic, prodromal and symptomatic carriers. Associations between longitudinal connectivity, and neurodegeneration and symptom severity highlight the potential of task-free fMRI for tracking disease progression during each clinical stage.

Future directions: This study lays important groundwork for tracking disease progression as early as the asymptomatic stage. Future research should establish phenotype- and stage-specific connectivity trajectories in carriers who convert to advanced stages or develop distinct *C9orf72*-associated syndromes.

## 1 Background

Understanding the earliest systems-level changes in genetic frontotemporal dementia (FTD) is a critical step toward forecasting symptom onset and tracking disease progression. Intrinsic connectivity networks (ICNs), as measured by task-free functional MRI (tf-fMRI), show distinct abnormalities in genetic frontotemporal lobar degeneration (FTLD), even during the asymptomatic stage when atrophy is subtle or absent [1–5]. The most common FTLD genetic variant, a hexanucleotide repeat expansion in *C9orf72,* most often presents with behavioral variant frontotemporal dementia (bvFTD), ALS, or bvFTD with motor neuron disease (bvFTD-MND) [6,7]. Most *C9orf72* tf-fMRI connectivity studies to date have been limited by cross-sectional designs, with small sample sizes, and/or a focus on a single clinical stage [2,8–15].

Cross-sectional studies have identified ICN alterations in *C9orf72* expansion carriers with bvFTD (*C9orf72*-bvFTD), ALS (*C9orf72-*ALS), or FTD-MND [8–15]. In a previous study of 14 *C9orf72-*bvFTD with or without MND [8], connectivity disruptions (i.e., reduced connectivity compared to controls) were identified in the salience network (SN), which is involved in processing emotionally salient stimuli [16], default mode network (DMN), associated with memory and meta-cognitive functions [17,18], and sensorimotor network (SMN), essential for motor functioning [19]. These ICNs have also been implicated in sporadic bvFTD and MND [20,21]. Additionally, *C9orf72*-bvFTD demonstrated ICN disruption to the medial pulvinar nucleus of the thalamus [8], an association nucleus involved in social cognition and in guiding visual attention toward salient stimuli [22]. Asymptomatic *C9orf72* expansion carriers also show functional alterations in these same networks [2,9,11], appearing decades before expected symptom onset and in the absence of substantial atrophy. In contrast to these early connectivity anomalies, gray matter (GM) reductions in asymptomatic carriers are subtle, with atrophy typically becoming evident during the symptomatic stage [14,23–30].

Longitudinal characterization of functional connectivity along various clinical stages of *C9orf72* would provide the foundational next step toward studies that anticipate symptom onset and monitor disease progression. Most longitudinal studies have included limited sample sizes that focus on a single clinical stage, and to date, no previous studies have examined longitudinal network connectivity in prodromal *C9orf72* or *C9orf72-*bvFTD. Longitudinal studies of *C9orf72*-ALS have reported a lack of connectivity changes compared to controls [13,24], possibly due to limited follow-up time and sample sizes. For asymptomatic carriers, two tf-fMRI studies involving the same 15 participants were conducted over a period of up to 18 months. One study identified regions of declining and increasing connectivity within a motor network and increasing connectivity within a speech production network in asymptomatic carriers compared to controls [9]. The other study found that asymptomatic carriers had increasing intra-network homogeneity in the somatomotor network [11]. Overall, these studies suggest that functional network abnormalities and their progressive alterations precede detectable structural degeneration, but the extent to which longitudinal connectivity changes appear in each clinical stage of *C9orf72* remains unclear.

In the present study, we analyzed longitudinal tf-fMRI and structural MRI changes in a cohort of 82 *C9orf72* expansion carriers [36 asymptomatic (aSxC9), 17 prodromal (proC9), and 29 symptomatic (SxC9)]. We aimed to examine longitudinal functional and structural changes for each clinical stage and to determine how imaging measures relate to symptom severity and plasma neurofilament light chain (NfL) concentration, a non-specific biomarker of axonal injury that increases around symptom onset in genetic FTD [31]. Using a seed-based connectivity approach, we examined the SN, DMN, SMN and a medial pulvinar thalamus network (MPN), networks which have shown alterations in cross-sectional studies [2,8]. We hypothesized that all three clinical stages would show longitudinal connectivity changes within these ICNs, but that longitudinal gray matter (GM) loss would be detectable only during the prodromal and symptomatic stages. We also hypothesized that longitudinal changes in connectivity or GM would correlate with baseline NfL and symptom severity in those participants with available data.

## 2 Methods

### 2.1 Participants

Participants were recruited from the University of California, San Francisco (UCSF) Memory and Aging center and the ARTFL/LEFFTDS Longitudinal Frontotemporal Lobar Degeneration (ALLFTD) study, a multisite genetic frontotemporal dementia study [32]. Clinical diagnoses were rendered at each study site.

We identified 82 *C9orf72* expansion carriers with usable neuroimaging data (Table 1). A pathological expansion was defined as having more than 40–50 repeats following previous methods [33], and all individuals tested negative for microtubule-associated protein tau (*MAPT*) [34] and progranulin (*GRN*) [35] variants. *C9orf72* expansion carriers were grouped based on the clinical diagnosis at the time of the first visit.

**Table 1:**
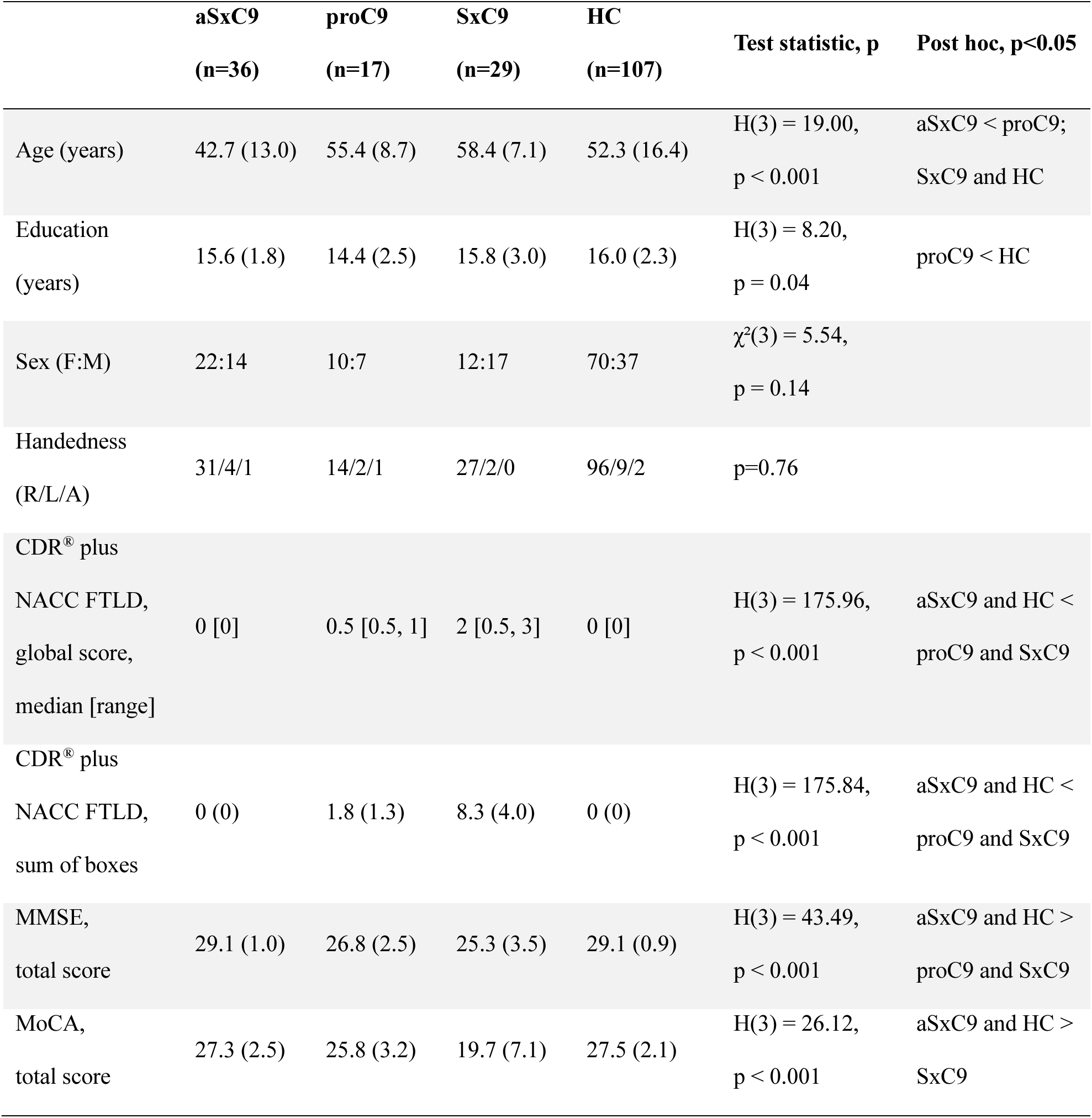
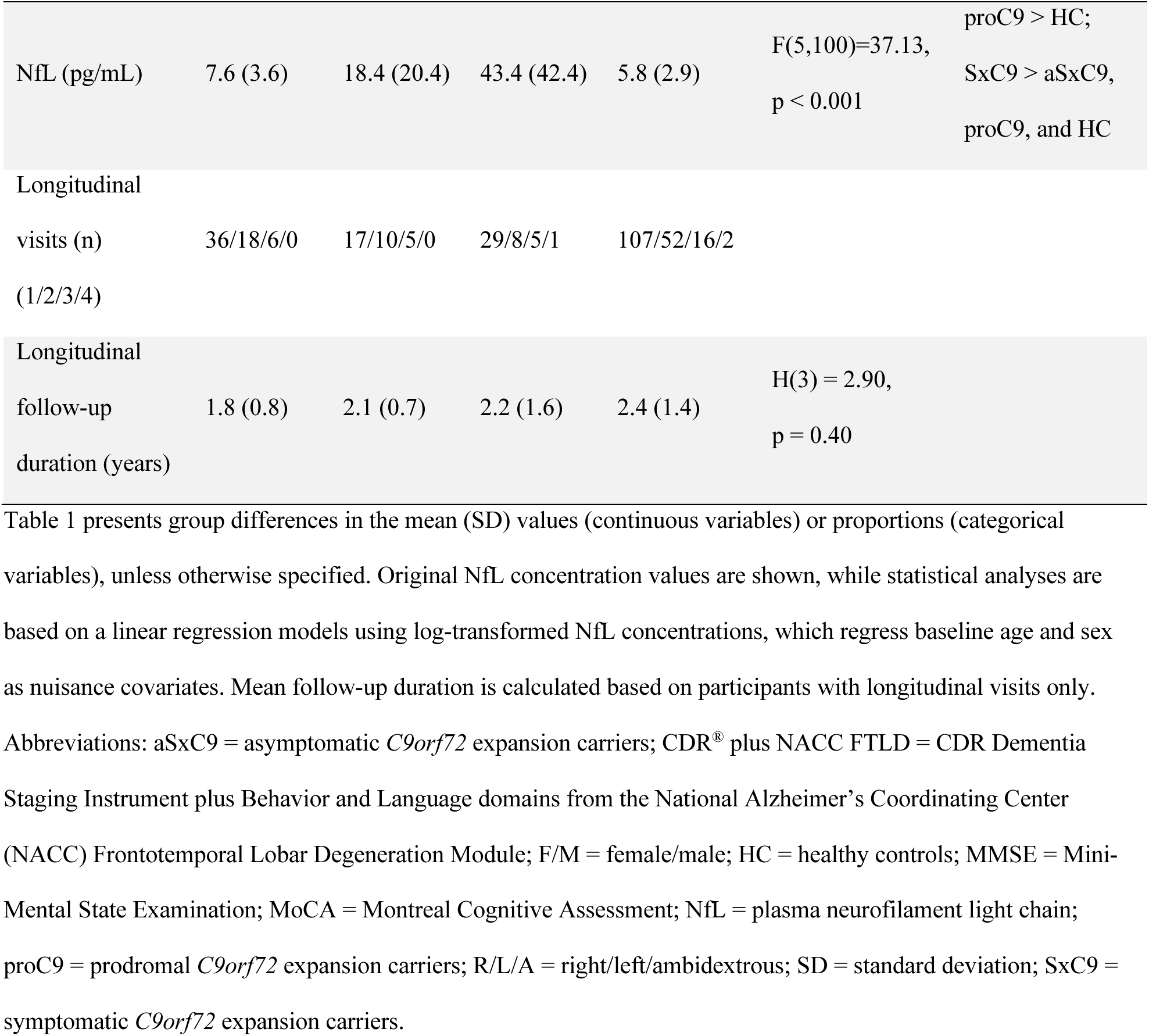
Baseline demographics and clinical measures of *C9orf72* expansion carriers and healthy controls.

The 36 aSxC9 were diagnosed as clinically normal and had a global score of 0 on the CDR plus Behavior and Language domains from the National Alzheimer’s Coordinating Center (NACC) FTLD module (CDR® plus NACC FTLD) [36]. The 17 proC9 included 15 diagnosed with mild cognitive impairment (MCI) [37], of whom two had a global score of 1 on the CDR® plus NACC FTLD: one with a mood disorder, and one with alcohol use. The 29 SxC9 included 17 bvFTD, 7 bvFTD-MND, one bvFTD with primary progressive aphasia of unspecified subtype, one ALS-MCI, and three ALS. To enable comparisons of connectivity abnormalities between each of the three *C9* cohorts and a common reference group, we included a single group of 107 healthy control participants (HC). HC consisted of noncarrier members of *GRN, MAPT*, and *C9orf72* families with unrelated controls who were included as necessary to ensure an age distribution comparable to the *C9orf72* expansion carriers. All HC were required to have a Mini-Mental State Examination (MMSE) score ≥ 27, and if unavailable, a corresponding score ≥ 21 on the Montreal Cognitive Assessment (MoCA) [38], a global score of 0 on the CDR® plus NACC FTLD, and no significant white matter lesions or history of neurological disease. Baseline MRI scanning was required to be within 180 days of clinical assessment.

### 2.2 Statistical analyses of demographic, clinical and neuropsychological measures

Demographic, clinical, and neuropsychological measures were compared between groups using one-way ANOVA or Kruskal-Wallis tests if the assumptions required for ANOVA were not met, implemented in R (version 4.3.1). Significant differences were followed by post hoc multiple comparisons: Tukey’s HSD after ANOVA or Dunn’s test (FSA v0.9.5) with Bonferroni adjustment after Kruskal-Wallis tests, with a significance threshold of p<0.05. For categorical variables, chi-square tests were used to compare groups when the cell frequencies were 5 or more, and Fisher’s exact tests when the cell frequencies were less than 5. A linear regression model was used to examine the effects of group on log-transformed NfL levels at baseline while controlling for baseline age and sex. Significant differences were followed by Tukey’s HSD post-hoc tests (p<0.05). Linear mixed effects (LME; lme4 v1.1.34) models were used to compare changes in clinical and neuropsychological measures over time between groups, controlling for baseline age, sex and education. HC was set as the reference group. The fixed effects included group, time since baseline, the interaction between group and time, and baseline age, sex and education. Random intercepts for each subject were included to account for repeated measures.

For measures for which a group had fewer than seven subjects with longitudinal data, that group was excluded from the LME analyses. To aid the interpretation of significant longitudinal group differences, raw annualized change for each outcome measure was calculated. For each subject, the difference between the last and first observed score was divided by the time (in years) between those visits to obtain an annualized rate of change. Group-level summaries were then computed as the mean and standard deviation of these per-subject annualized changes.

### 2.3 Image acquisition

Each participant underwent a T1-weighted (T1w) structural MRI scan and a T2*-weighted echo-planar imaging tf-fMRI scan on a 3T scanner at each site. The tf-fMRI session acquired either 240 (8:08 minutes) or 197 volumes (10 minutes), following previous protocols at the UCSF Memory and Aging Center and ALLFTD [39].

Image quality was assessed for protocol compliance, excessive motion and other image artifacts. Quantitative fMRI motion criteria required that participants had a mean framewise displacement (FD) below 0.5 mm and at least 4 minutes of data meeting this threshold. Voxel-based morphometry analyses and functional imaging were conducted using T1w and tf-fMRI scans acquired at the same visit.

### 2.4 Voxel-based morphometry analysis

#### 2.4.1 Preprocessing

T1w images were processed with the standard longitudinal voxel-based morphometry pipeline in SPM12 (Wellcome Trust Centre for Neuroimaging, UCL, London, UK) embedded in MATLAB (version R2021a). Structural T1w images of each participant at each visit were bias field corrected and segmented into tissue probability maps using the “New Segment” routine [40]. The tissue maps were normalized to standard MNI space using serial longitudinal registration, via a subject-specific average template created using non-linear diffeomorphic and rigid-body registration [41]. Next, the maps were modulated by multiplying the timepoints’ Jacobians with the corresponding tissue maps, and smoothed using an 8 mm FWHM isotropic Gaussian kernel. Finally, these GM maps were parcellated into 246 regions of interest (ROIs) based on the Brainnetome atlas [42], and gray matter volume (GMV) within each ROI was obtained for subsequent statistical analyses. Total intracranial volume (TIV) was estimated by the sum of GM, white matter, and cerebrospinal fluid volumes.

#### 2.4.2 Harmonization

To harmonize multisite structural data for subsequent statistical analyses, a customized ComBat approach [4] was applied to individual GM maps to remove batch effects of no interest (different scanners), while preserving biological variability of interest (i.e., age, sex, education and handedness). ComBat parameters were estimated in an independent group of 83 HC (Combat HC; Supplementary Table 1) whose demographic characteristics spanned the age ranges and scanner sites of the study cohorts and were used to harmonize study cohort data.

#### 2.4.3 Statistics

Longitudinal changes in GMV were examined using LME models (lme4 v1.1.34, lmerTest v3.1.3), with fixed effects for group, time between baseline and follow-up, group-by-time interaction, age, sex, education, and TIV. Random effects included individual intercepts. Bonferroni correction (p<0.05) was applied for the 246 ROIs.

### 2.5 Functional imaging analysis

#### 2.5.1 Preprocessing

Data were preprocessed using the fMRIPrep pipeline [43]. Each T1w image was corrected for intensity non-uniformity (N4BiasFieldCorrection, ANTs v2.2.0), and used as the T1w-reference throughout the workflow. The T1w-reference was then skull-stripped, segmented to GM, white matter, and cerebrospinal fluid (fast, FSL v5.0.9). Volume-based spatial normalization to one standard space was performed through first creating an unbiased subject template by averaging T1w acquired across the visits included in the analysis, and then normalized to the template (FSL’s MNI ICBM 152 non-linear 6th Generation Asymmetric Average Brain Stereotaxic Registration Model; antsRegistration, ANTs 2.2.0), using brain-extracted versions of both T1w-reference and the template.

The first five fMRI volumes were dropped for magnetic field stabilization, and the following preprocessing was performed. First, a reference volume and its skull-stripped version were generated using a custom methodology of fMRIPrep. The blood-oxygen-level-dependent (BOLD) reference was then co-registered to the T1w-reference using boundary-based registration (bbregister, FreeSurfer v6.0.0) with six degrees of freedom. Head-motion parameters with respect to the BOLD reference (transformation matrices, and six corresponding rotation and translation parameters) were estimated before spatiotemporal filtering (mcflirt, FSL v5.0.9).

BOLD data were slice-time corrected (3dTshift, AFNI v16.02.07). BOLD time series were resampled onto their original, native space by applying a single, composite transform to correct for head-motion and susceptibility distortions. To normalize BOLD time-series into MNI152 standard space, motion correcting transformations, BOLD-to-T1w transformation and T1w-to-MNI warp were concatenated and applied in a single step (antsApplyTransforms, ANTs v2.1.0; Lanczos interpolation). Data were smoothed with a 6 mm full width at half maximum Gaussian kernel (SUSAN) [44]. Following these steps, we applied “non-aggressive” ICA-AROMA [45] to identify independent components representing motion-related artifacts based on spatial and temporal discriminative features, and regressed them from the time series. Finally, data underwent linear detrending, band-pass filtering (0.008 ∼ 0.08 Hz), and nuisance regression (the six head motion parameters, six head motion parameters one time point before, and the 12 corresponding squared items, mean signals from the white matter and cerebrospinal fluid, and global signal).

#### 2.5.2 Seed-based connectivity maps

We used 4 mm-radius spheres (i.e., seed regions) around peak coordinates from the literature (Supplementary Table 2) [2,8] to derive ICNs that show alterations in asymptomatic and symptomatic *C9orf72* expansion carriers cross-sectionally, including the SN, SMN, DMN and MPN. We conducted separate bivariate regression analyses between the average time series within each seed region and time series in the remaining voxels across the whole brain for each ICN of interest. This resulted in four ICN maps for each participant.

#### 2.5.3 Harmonization

Similar to GM maps, ICN maps were harmonized with ComBat based on the same Combat HC group. Covariates included age, sex, education, and handedness.

#### 2.5.4 Statistics

Voxelwise analyses were carried out in R (version 4.3.3) using the neuropointillist toolbox (0.0.0.9, http://ibic.github.io/neuropointillist) [46] on each ICN map separately. To assess cross-sectional differences between groups (aSxC9, proC9, SxC9, HC), we analyzed participants’ first visits using GLM. These models evaluated group effects with nuisance covariates for age, sex, education, handedness, and eyes-open status during scan acquisition. Linear contrasts were used to compare mean group levels, and interaction contrast tests were conducted to compare: 1) aSxC9 and HC, 2) proC9 and HC, and 3) SxC9 and HC.

Longitudinal change patterns were assessed by LME models (lme4 v1.1-35.1, lmerTest v3.1-3) with fixed effects terms for group (aSxC9, proC9, SxC9, HC), age at baseline, time since baseline, sex, education, handedness, and eyes-open status during scan acquisition. A random intercept was included for subject identity. LME models were fitted using restricted maximum likelihood (ReML). All available sessions were included to maximize the estimation accuracy of between-subject variability. For further analysis, results were converted to *Z* scores using Satterthwaite approximation to estimate the effective degrees of freedom. Based on the omnibus model with all four subgroups, the marginal means of linear trends (group × time since baseline interaction; emmeans, v1.9.0) associated with each subgroup were compared using planned linear contrasts and interaction contrasts. These tests compared 1) aSxC9 and HC, 2) proC9 and HC, and 3) sxC9 and HC.

A follow-up analysis explored the relationship between baseline CDR^®^ plus NACC FTLD sum of boxes [36] and longitudinal connectivity changes in proC9 (n=10) and SxC9 (n=8) separately using LME models, with fixed-effects terms for CDR^®^ plus NACC FTLD sum of boxes at baseline, baseline age, time since baseline, sex, education, handedness, and eyes-open status. A random intercept was included for subject identity.

All connectivity analyses were masked to the relevant network (Supplementary Figure 1). Network masks were created by using 1-sample t-tests of seed-based ICN maps in an independent group of 103 HC (Seed mask HC; Supplementary Table 1), thresholded at t ≥ 4. Results were family-wise error (FWE) corrected based on Monte-Carlo simulations using AFNI’s 3dClustSim program (NN1) [47]. Spatial smoothness was estimated for each subject and network using 3dFWHMx [48] based on data that was smoothed and detrended but not denoised by ICA-AROMA, regression, or temporal filtering. The spherical autocorrelation function parameters were averaged across all scans in the analysis when simulating the noise distribution. Significance was set at voxelwise thresholds of p < 0.05, with a cluster-level FWE-correction applied at p<0.05 using extent thresholding. We used one-tailed inference for linear contrasts for consistency with our previous findings [2–4].

### 2.6 Longitudinal changes associated with NfL concentrations

Plasma NfL concentrations were assayed utilizing the Simoa NF-light Advantage Kit from Quanterix on a SiMoA HD-X Analyzer instrument following previous protocols [49–51] at two processing labs.

The relationships between baseline log(NfL) concentrations and longitudinal change patterns in GMV in 246 ROIs and voxel-wise functional connectivity were assessed using LME models within each group. LMEs were run for aSxC9 (n=16) and a combined cohort of proC9 (n=8) and SxC9 (n=6), due to limited baseline NfL data for proC9 and SxC9. The models included fixed-effects terms for log(NfL) at baseline, age at baseline, time since baseline, sex, education, handedness, TIV (GMV only) and eyes-open status (functional connectivity only). A random intercept was included for subject identity. Results were masked to the relevant network and considered significant at voxelwise thresholds of p<0.05, with cluster-level FWE-correction at p<0.05.

## 3 Results

### 3.1 Demographic and clinical features, longitudinal gray matter comparisons

Demographic, clinical, and neuropsychological measures were compared between groups at baseline (Table 1, 2) and longitudinally (Table 3). Groups differed in age and education, with aSxC9 being younger than all other groups, and proC9 having fewer years of education than HC. As expected, aSxC9 showed no differences in any clinical or neuropsychological measures compared to HC. SxC9 (vs. HC) had lower scores in most measures, except for the Multilingual Naming Test and for this test, Sx had lower scores compared to aSx. ProC9 had impairments in Trail Making Tests, Semantic Fluency Test, and Benson Figure Recall Test compared to HC. Detailed group comparison results are reported in Table 2.

**Table 2:**
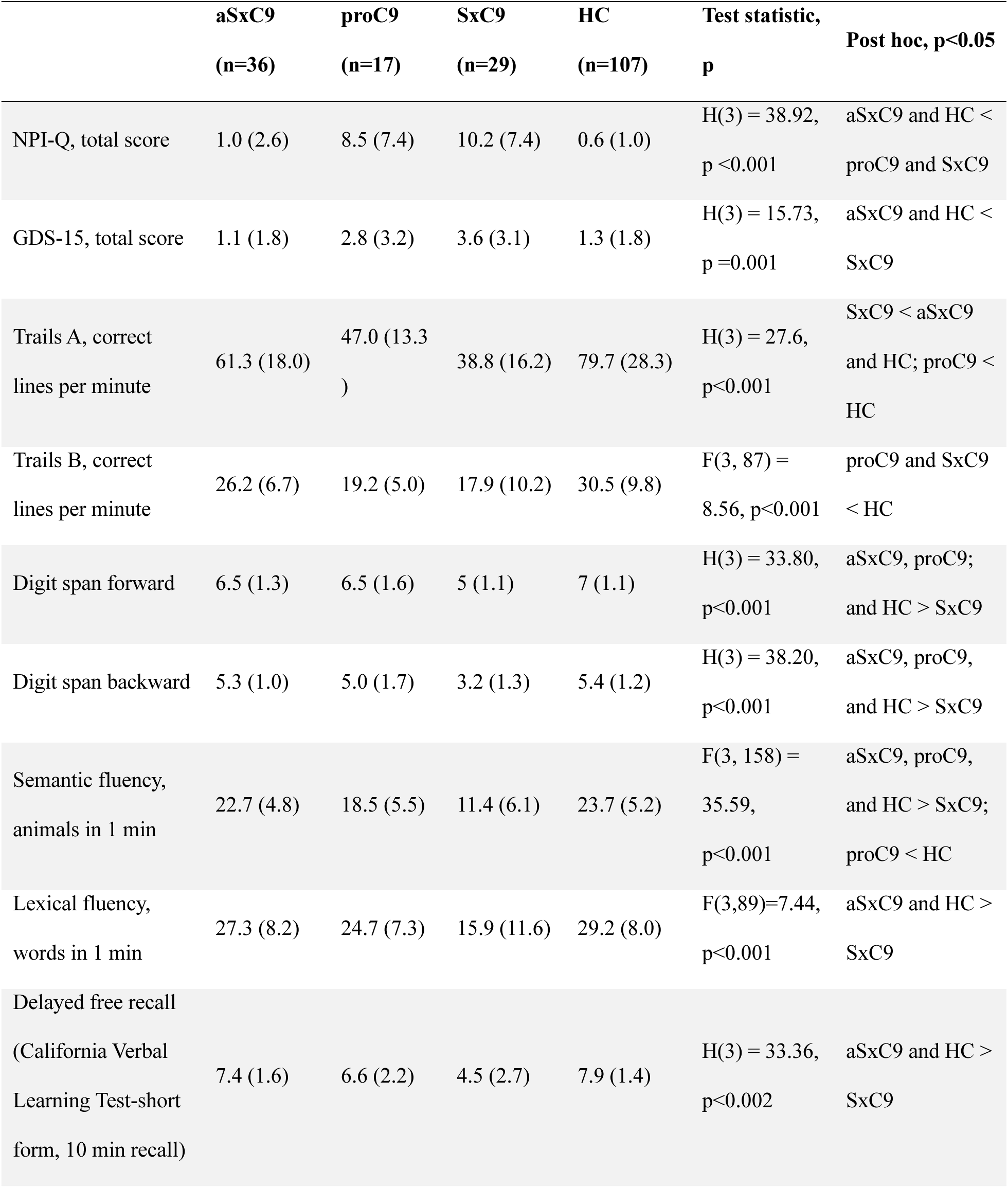
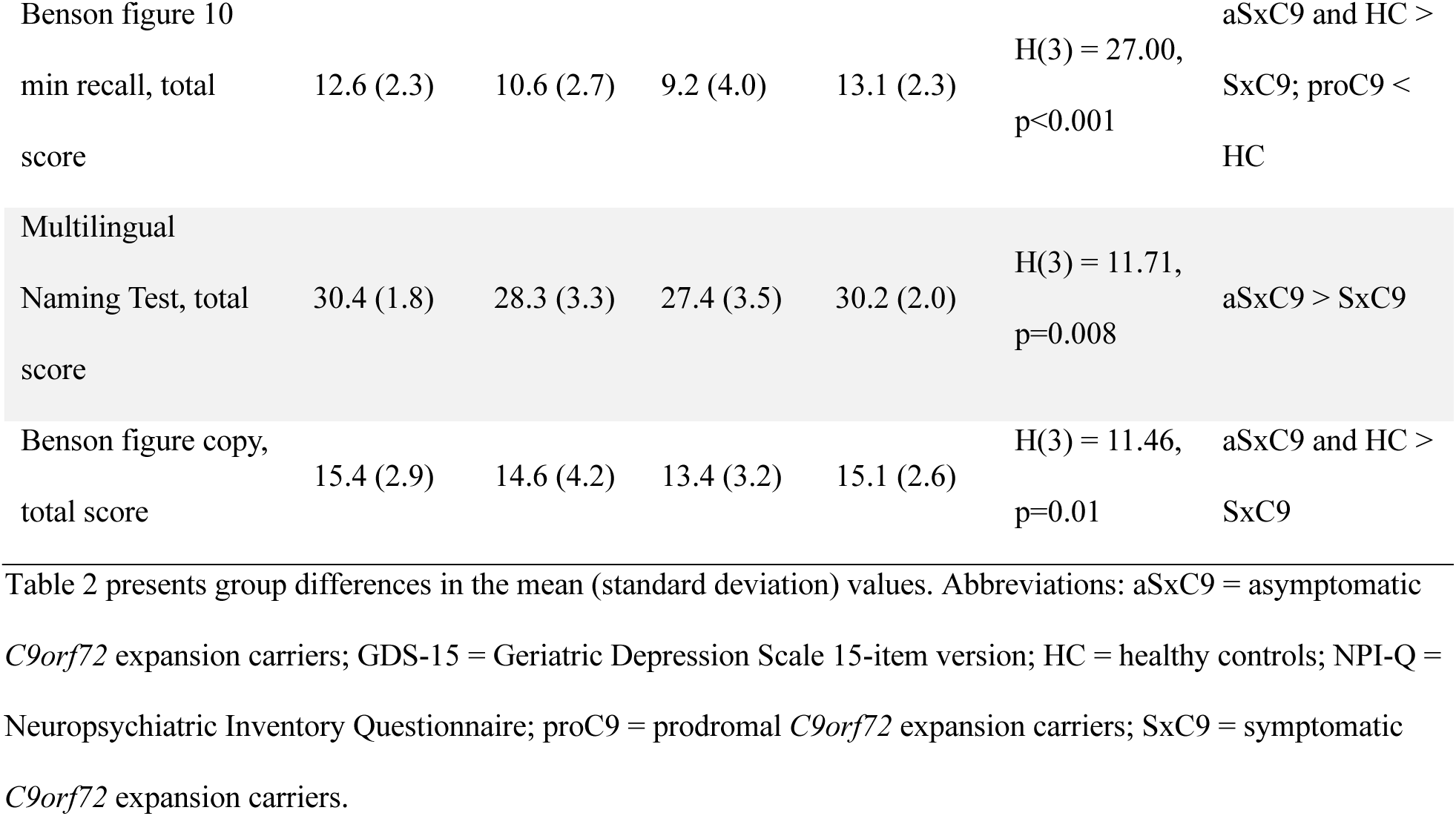
Baseline neuropsychological measures of *C9orf72* expansion carriers and healthy controls.

**Table 3:**
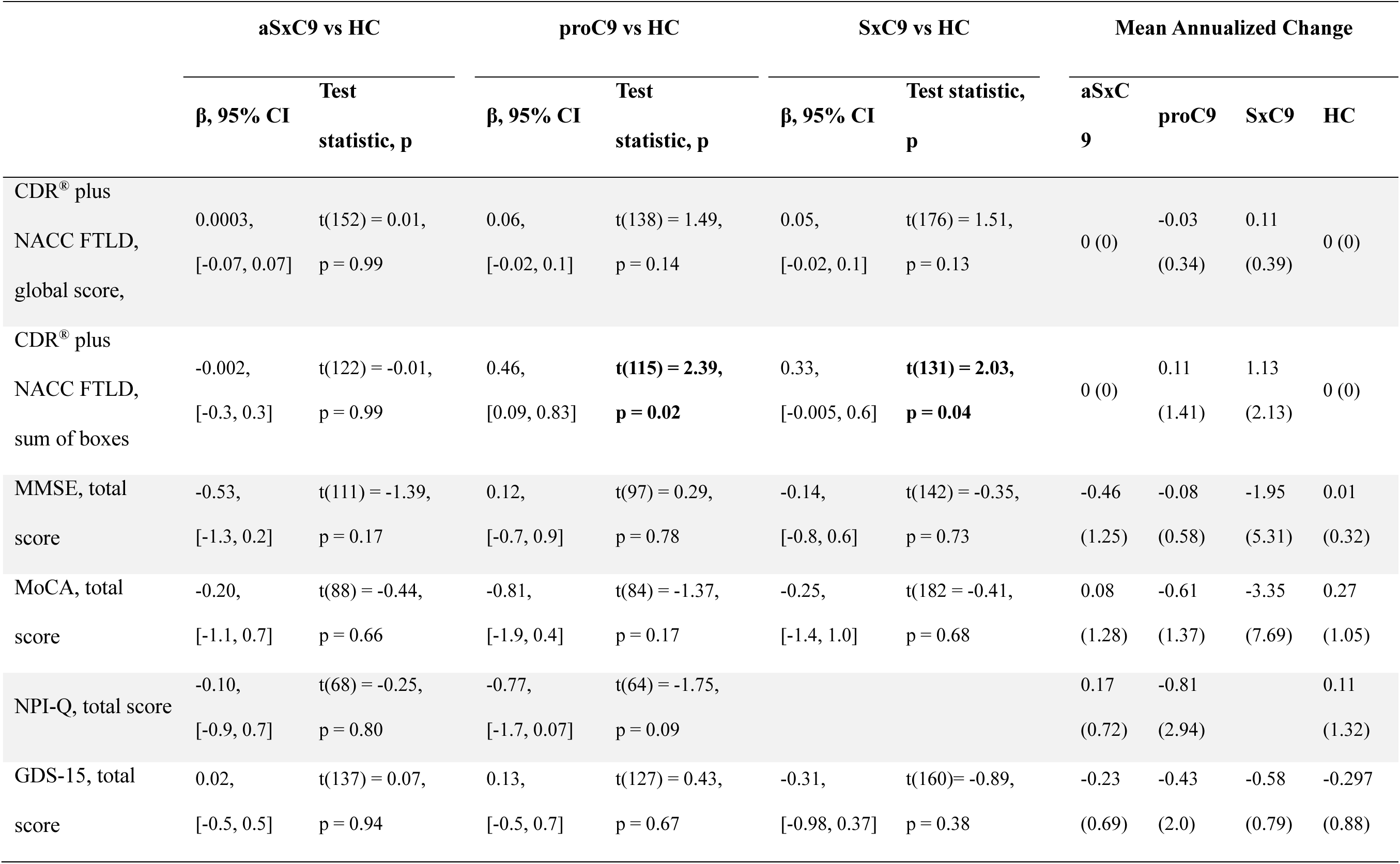
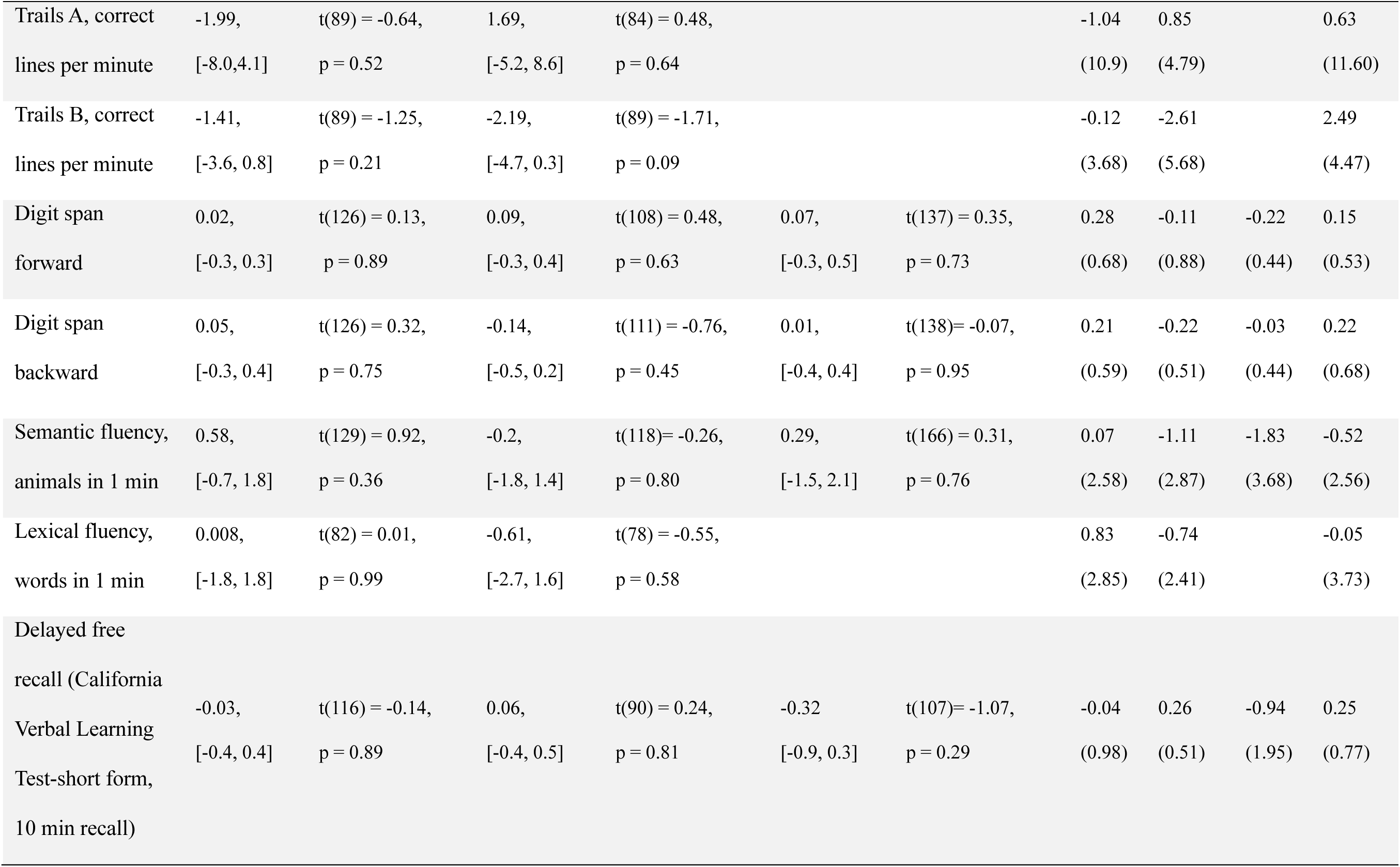
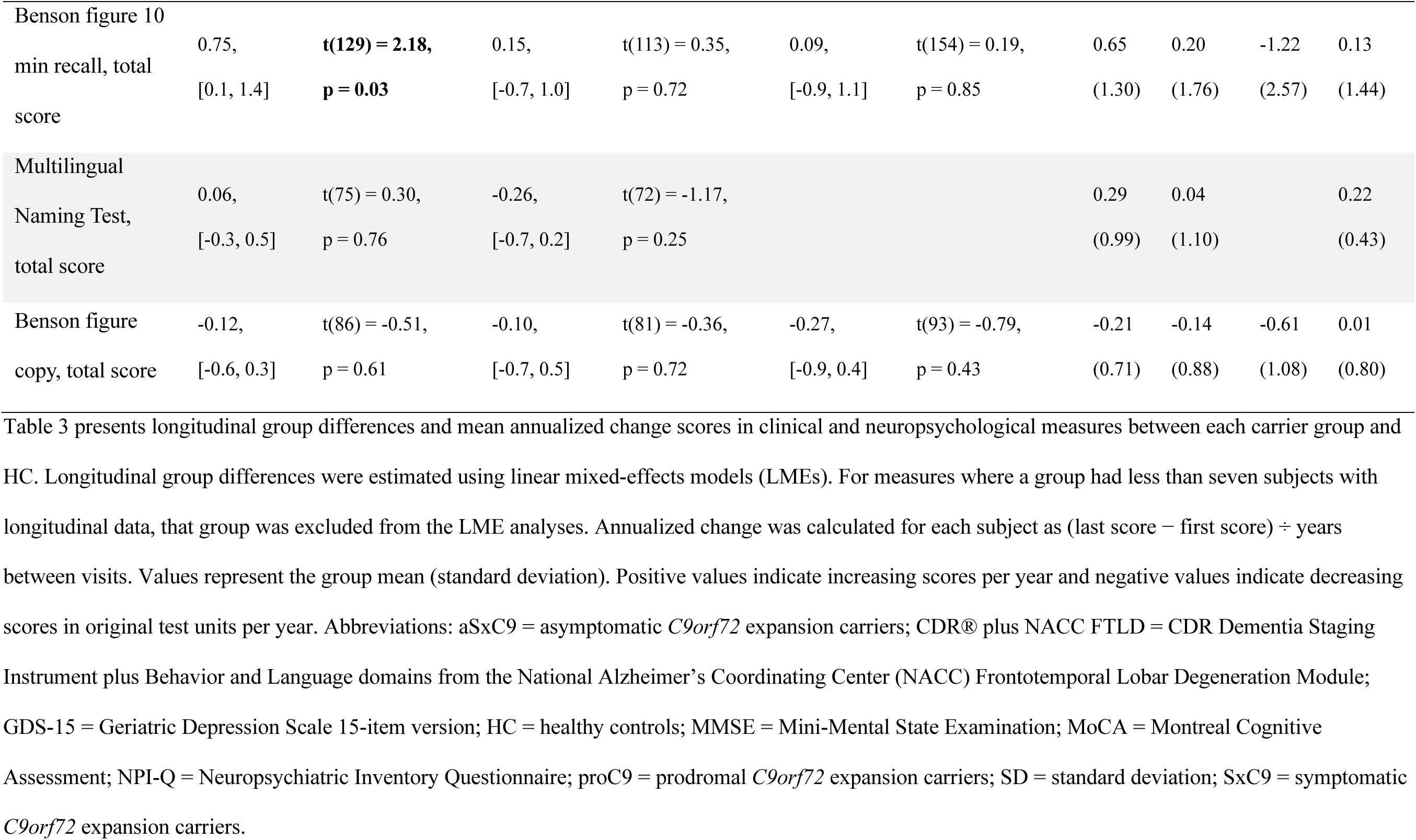
Longitudinal comparisons of clinical and neuropsychological measures in *C9orf72* expansion carriers and healthy controls.

Longitudinally, aSxC9 (vs. HC) showed slower decline in a visual memory task (Benson Figure Recall) compared to HC. Both proC9 and SxC9 demonstrated a faster rate of clinical decline (CDR^®^ plus NACC FTLD sum of boxes) compared to HC. There were no differences in longitudinal follow-up duration between the groups.

No carrier group demonstrated significant longitudinal GMV decline relative to HC (data not shown).

### 3.2 Asymptomatic carriers had baseline hypoconnectivity and longitudinal connectivity changes within *C9orf72*-relevant networks

At baseline, aSxC9 showed connectivity disruptions (i.e., lower connectivity) within networks previously reported to show alterations in *C9orf72* expansion carriers (Figure 1A). SN hypoconnectivity (vs. HC) was detected in the bilateral anterior cingulate and medial prefrontal cortex, left anterior insula and angular gyrus, right orbitofrontal cortex and middle temporal gyrus. Within the SMN, decreased connectivity was seen bilaterally in the primary motor cortex, supplementary motor area, and occipital lobe. MPN hypoconnectivity (vs. HC) was concentrated in bilateral posterior thalamus, precentral and postcentral gyrus, midcingulate, and precuneus. No significant alterations in DMN connectivity were found.

**Figure 1.**
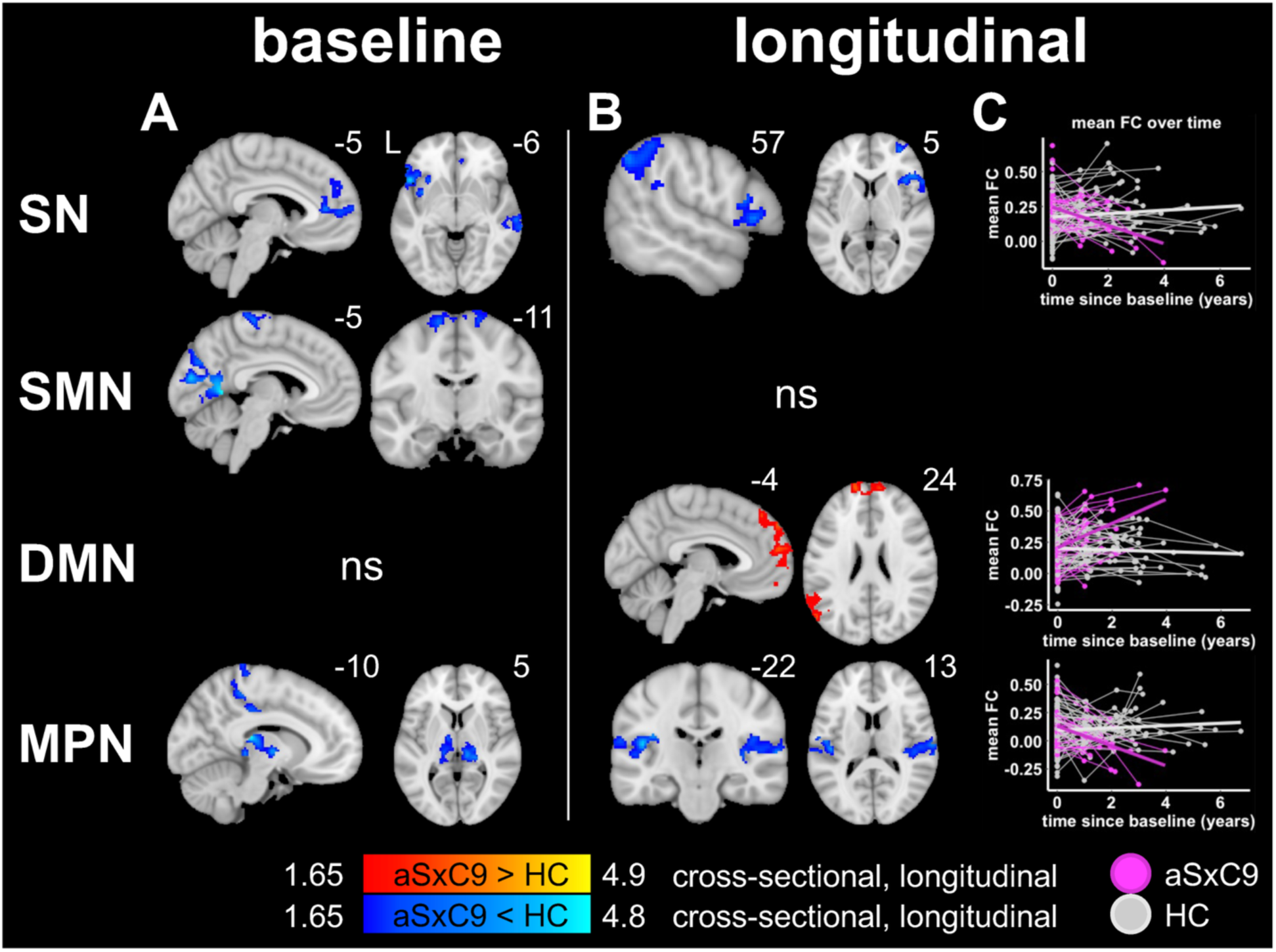
Asymptomatic carriers exhibit baseline hypoconnectivity and longitudinal connectivity changes within *C9orf72*-relevant networks. (A) At baseline, aSxC9 show regions of connectivity disruptions including in key regions of the SN (bilateral medial prefrontal cortex and left anterior insula), SMN (bilateral primary motor cortex and supplementary motor area), and MPN (bilateral posterior thalamus) compared to HC. (B) Longitudinally, aSxC9 show regions of connectivity declines in the SN and MPN, and increasing connectivity in the DMN. Notably, longitudinal SN connectivity decline is observed in the right anterior insula, which shows baseline connectivity disruption contralaterally. (C) Mean connectivity versus time since the baseline visit within the significant voxels shown in (B) is plotted for visualization purposes only. Results are thresholded at p<0.05 voxelwise, with cluster-level FWE correction at p<0.05, masked to the relevant network mask. Color bars represent z-scores for cross-sectional alterations or longitudinal changes, and result maps are superimposed on the MNI brain template. Abbreviations: aSxC9 = asymptomatic *C9orf72* expansion carriers; DMN = default mode network; FC = functional connectivity; HC = healthy controls; MPN = medial pulvinar network; ns = not significant; SMN = sensorimotor network; SN = salience network.

Longitudinally, aSxC9 showed declining connectivity in the SN and MPN compared to HC (Figure 1B-C). SN connectivity declines were observed in the right middle and superior frontal cortex, ventrolateral prefrontal cortex, anterior insula, and inferior parietal lobule. Of note, longitudinal SN connectivity decline arose in the right anterior insula. MPN connectivity declines emerged in bilateral superior temporal gyrus extending to the operculum. For the DMN, aSxC9 showed increasing connectivity principally in bilateral medial prefrontal cortex and left middle temporal gyrus and angular gyrus. No significant longitudinal changes in SMN connectivity were observed.

### 3.3 Prodromal carriers showed baseline connectivity alterations but lacked detectable longitudinal connectivity changes compared to HC

At baseline, proC9 had regions of SMN hypoconnectivity, and hyperconnectivity in the DMN and MPN compared to HC (Figure 2). Specifically, hypoconnectivity was observed in key regions of the SMN, including the right pre- and post-central gyrus, bilateral supplementary motor area, and occipital regions. DMN hyperconnectivity was detected in bilateral midcingulate cortex, precuneus, posterior cingulate, and occipital regions. MPN hyperconnectivity was seen in bilateral posterior cingulate and precuneus extending to the cuneus. ProC9 showed no detectable alterations in the SN, nor longitudinal connectivity changes in any networks compared to HC.

**Figure 2.**
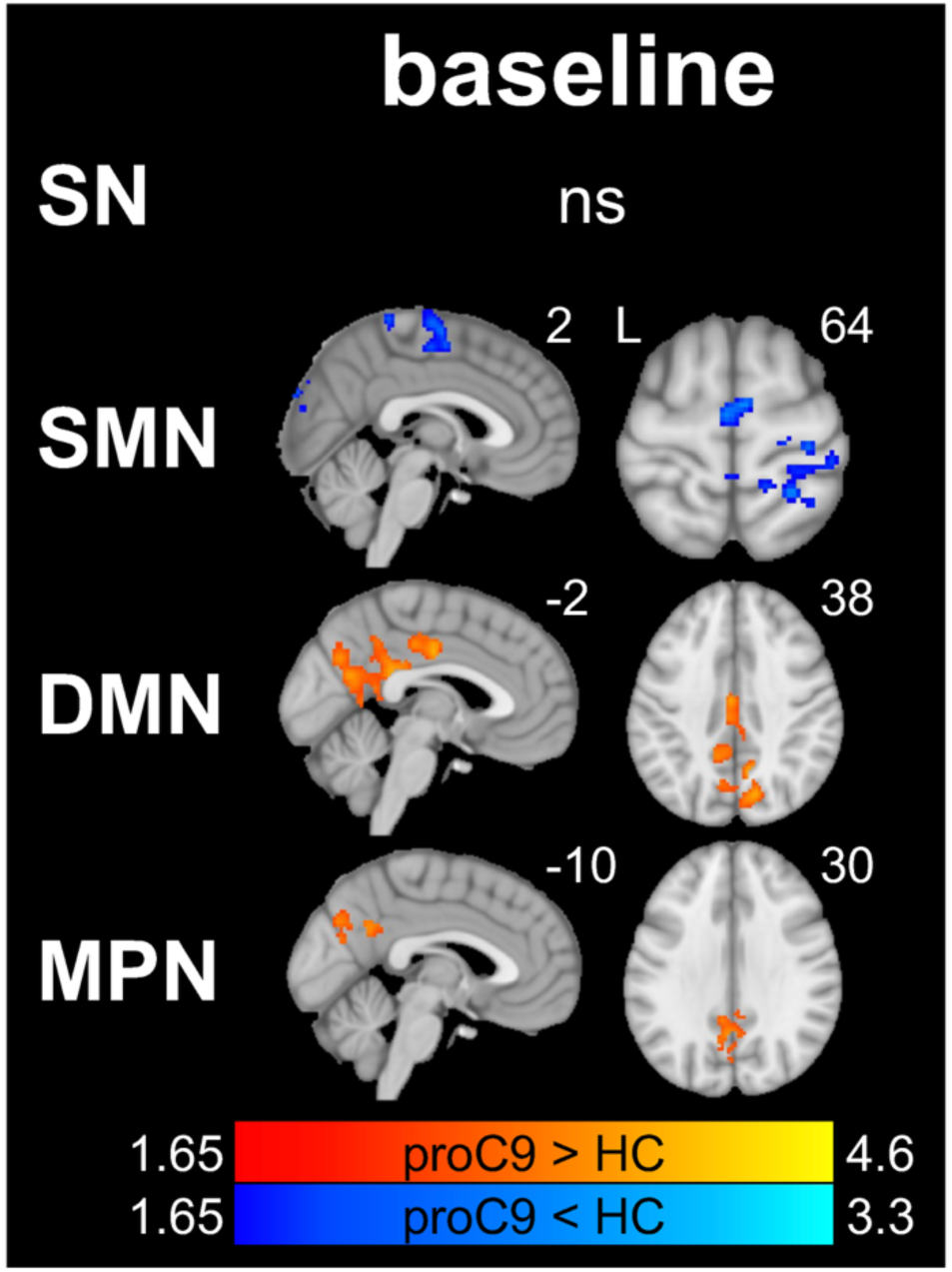
Group comparison of prodromal carriers versus HC identify baseline alterations in network connectivity, but a lack of detectable longitudinal connectivity changes. At baseline, proC9 display regions of SMN hypoconnectivity which include bilateral perirolandic regions, and bilateral posterior cingulate and parietal regions of hyperconnectivity in the DMN and MPN compared to HC. No significant longitudinal connectivity changes are observed. Results are thresholded at p<0.05 voxelwise, with cluster-level FWE correction at p<0.05, masked to the relevant network mask. Color bars represent z-scores, and result maps are superimposed on the MNI brain template. Abbreviations: DMN = default mode network; HC = healthy controls; MPN = medial pulvinar network; ns = not significant; proC9 = prodromal *C9orf72* expansion carriers; SMN = sensorimotor network; SN = salience network.

### 3.4 Symptomatic carriers had predominantly baseline hypoconnectivity and longitudinal increases within networks examined

SxC9 showed SN and SMN connectivity disruptions compared to HC (Figure 3A). Specifically, SN hypoconnectivity was identified in bilateral anterior cingulate, medial prefrontal cortex, right insula and ventrolateral prefrontal cortex. SMN hypoconnectivity was found in bilateral perirolandic cortex and supplementary motor area. For the DMN, divergent connectivity alterations included hypoconnectivity in regions of right superior and middle frontal cortex, accompanied by hyperconnectivity in bilateral mid/posterior cingulate, precuneus, and left middle temporal gyrus. No significant connectivity alterations were detected in the MPN at baseline.

**Figure 3.**
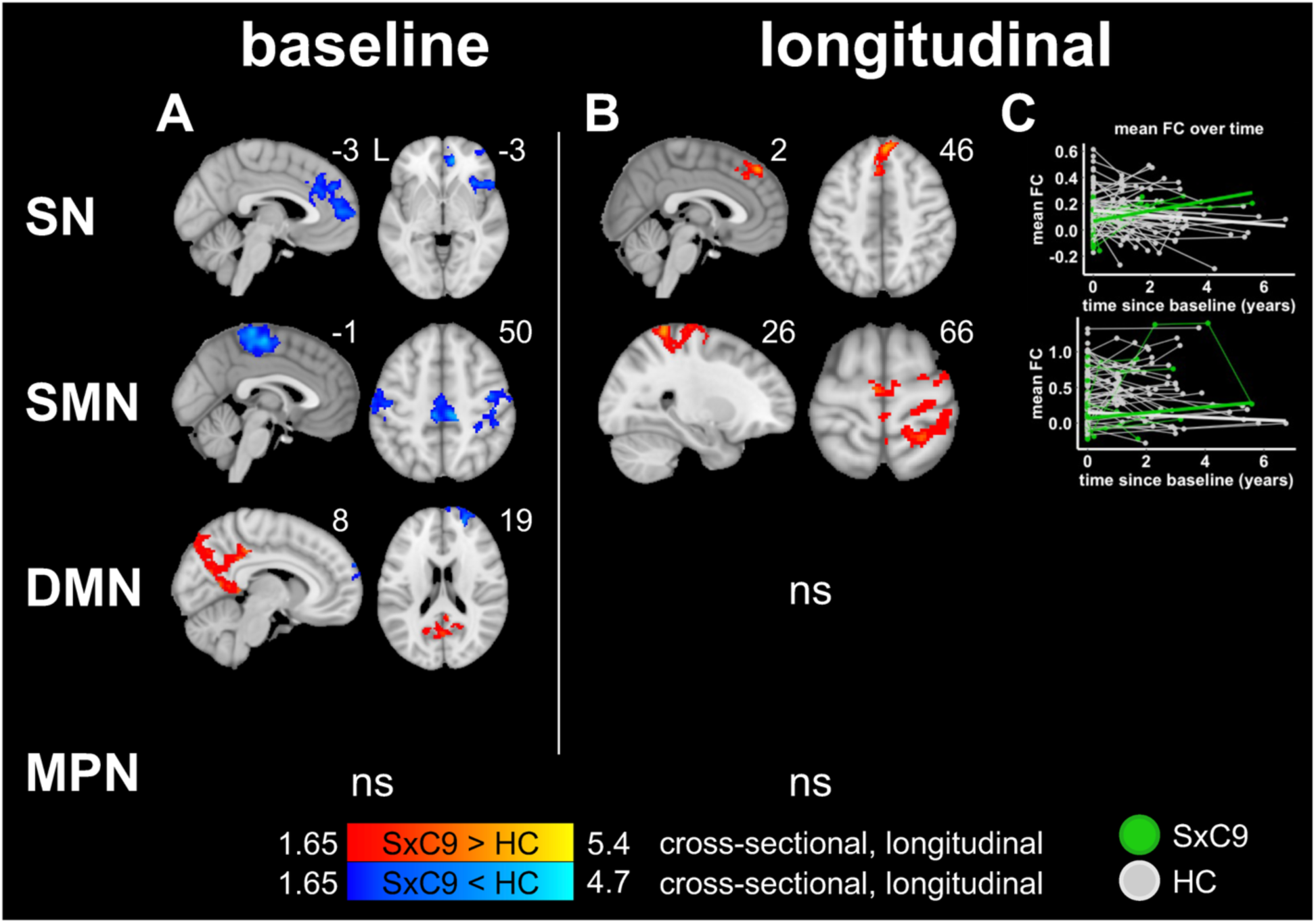
Symptomatic carriers have baseline connectivity alterations and longitudinal connectivity increases in the SN and SMN. (A) At baseline, SxC9 (vs. HC) show connectivity disruptions in the SN and SMN, and divergent connectivity alterations in the DMN. (B) At follow-up, SxC9 exhibit connectivity increases over time within the SN in the bilateral superior medial prefrontal cortex, and within the SMN in bilateral perirolandic regions. These changes overlap well with regions showing baseline connectivity disruptions in the same networks. (C) Mean connectivity versus time since the baseline visit within the significant voxels shown in (B) is plotted for visualization purposes only. Results are thresholded at p<0.05 voxelwise, with cluster-level FWE correction at p<0.05, masked to the relevant network mask. Color bars represent z-scores for cross-sectional alterations or longitudinal changes, and result maps are superimposed on the MNI brain template. Abbreviations: DMN = default mode network; FC = functional connectivity; HC = healthy controls; MPN = medial pulvinar network; ns = not significant; SMN = sensorimotor network; SN = salience network; SxC9 = symptomatic *C9orf72* expansion carriers.

SxC9 demonstrated longitudinal increases in the SN and SMN (Figure 3B). Increasing SN connectivity appeared in bilateral superior medial prefrontal cortex, while increasing SMN connectivity emerged in bilateral perirolandic regions. Notably, these SN and SMN regions showing longitudinal increases overlapped well with regions showing connectivity disruptions at baseline. No significant longitudinal changes were detected in the DMN and MPN.

### 3.5 Higher baseline NfL was associated with longitudinal connectivity changes in aSxC9, and longitudinal DMN increases and GMV decline in prodromal/symptomatic carriers

For aSxC9, baseline NfL concentrations showed an association with longitudinal connectivity changes in the SMN and MPN (Figure 4A). Higher NfL concentrations were associated with longitudinal SMN connectivity decreases within the right postcentral gyrus. Additionally, higher NfL concentrations were associated with longitudinal MPN connectivity increases in the right anterior thalamus as well as longitudinal decreases in this network in the bilateral posterior cingulate, precuneus, and left middle temporal gyrus. No significant associations were found between baseline NfL concentrations and longitudinal GMV changes, nor connectivity changes in the SN or DMN for aSxC9.

**Figure 4.**
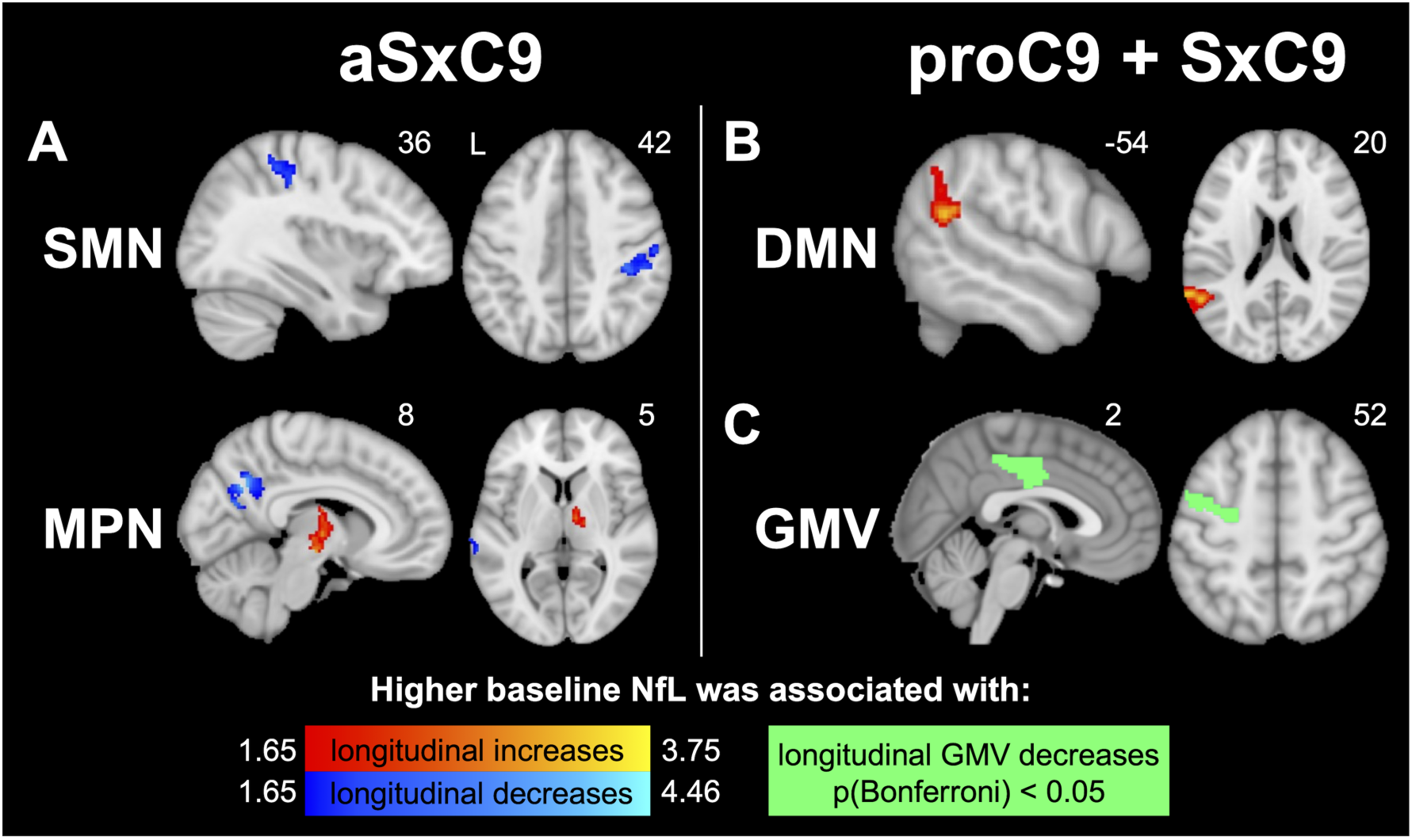
Baseline NfL concentrations are associated with longitudinal GMV declines and functional connectivity changes in *C9orf72* expansion carriers. (A) In aSxC9, greater NfL concentrations at baseline are associated with longitudinal SMN connectivity decline in a right postcentral gyrus region, and both connectivity decreases (bilateral parietal and left temporal regions) and increases (right anterior thalamus) in the MPN. (B) A combined group of proC9 (n=8) and SxC9 (n=6) show that higher baseline NfL concentrations correlate with longitudinal DMN increases in the left angular gyrus and longitudinal atrophy in the left precentral gyrus and right midcingulate cortex. Connectivity results are thresholded at p<0.05 voxelwise, with cluster-level FWE correction at p<0.05, masked to the relevant network mask. Color bars represent z-scores for association analyses with longitudinal connectivity, or indicate regions with NfL-associated GMV declines surviving multiple comparisons correction. Result maps are superimposed on the MNI brain template. Abbreviations: aSxC9 = asymptomatic *C9orf72* expansion carriers; DMN = default mode network; FC = functional connectivity; GMV = gray matter volume; MPN = medial pulvinar network; NfL = plasma neurofilament light chain; proC9 = prodromal *C9orf72* expansion carriers; SMN = sensorimotor network; SxC9 = symptomatic *C9orf72* expansion carriers.

Given the limited NfL data for proC9 (n=8) and SxC9 (n=6) participants at baseline, we combined these groups for our analyses. In this proC9 and SxC9 combined group, higher baseline NfL concentrations were associated with longitudinal DMN connectivity increases within the left angular gyrus (Figure 4B). Higher baseline NfL concentrations were also associated with longitudinal GMV atrophy in the left precentral gyrus and right midcingulate cortex (Figure 4C). No significant associations were observed between baseline NfL concentrations and longitudinal connectivity changes in the SN, SMN, or MPN for this group.

### 3.6 In prodromal and symptomatic carriers, longitudinal connectivity changes in the SN and SMN were associated with greater baseline symptom severity

Although proC9 lacked longitudinal connectivity changes (Figure 2) over the relatively limited average follow up of 2.1 years, we identified longitudinal connectivity changes in the SN and SMN that were associated with a measure of symptom severity (Figure 5A). Greater symptom severity (higher baseline CDR^®^ plus NACC FTLD sum of boxes scores) correlated with longitudinal SN connectivity declines in medial frontal and anterior cingulate cortex, and with longitudinal SMN increases in bilateral precentral gyrus.

**Figure 5.**
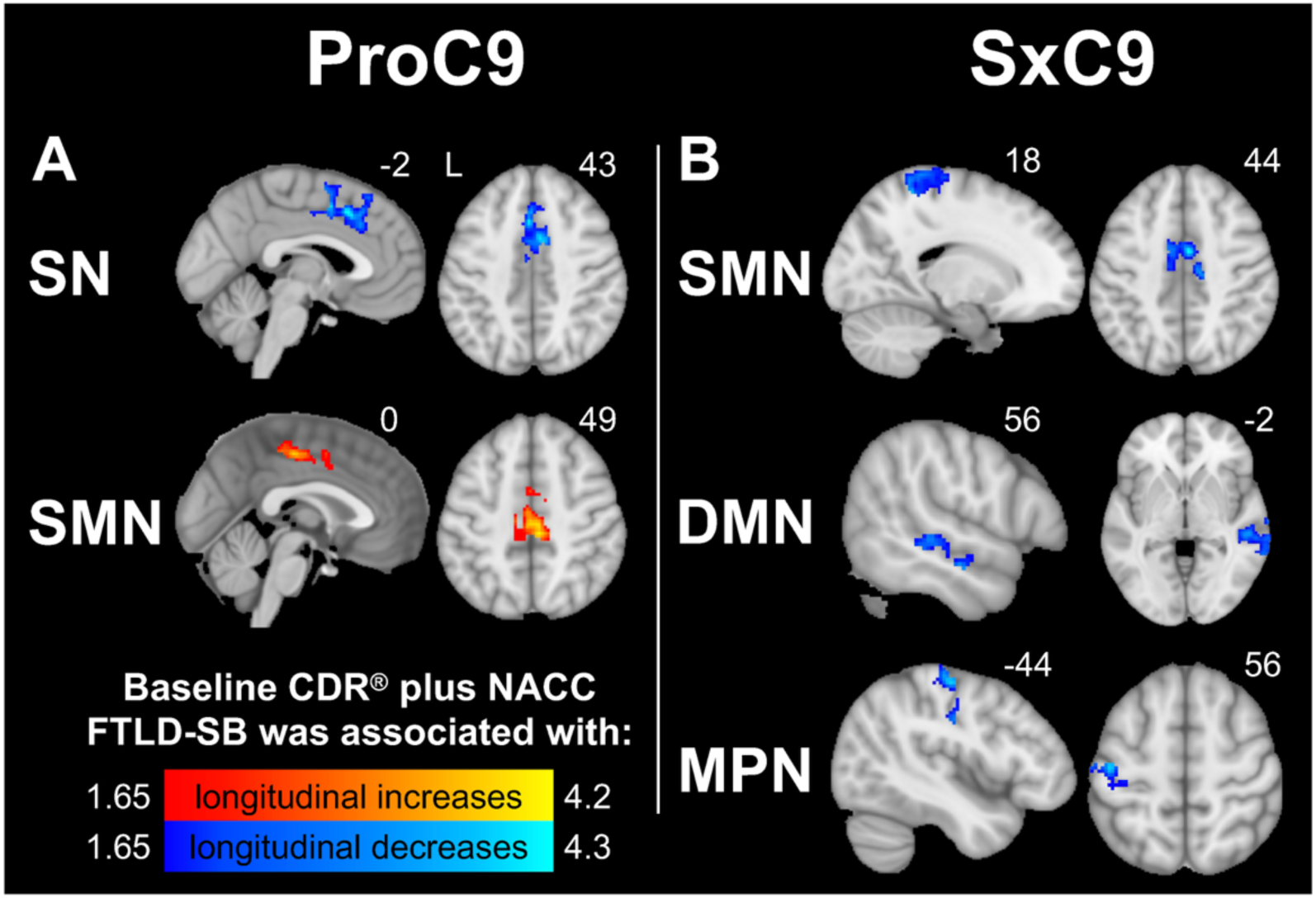
In prodromal and symptomatic carriers, greater symptom severity is associated with longitudinal connectivity changes. (A) In proC9, greater symptom severity (higher FTLD-CDR-SB scores) are associated with longitudinal SN decreases in the bilateral medial prefrontal cortex and anterior cingulate cortex, and SMN increases in bilateral supplementary motor areas. (B) In SxC9, higher scores correlate with longitudinal declines in the SMN (right perirolandic regions and bilateral midcingulate cortex), DMN (right middle temporal gyrus) and MPN (left perirolandic regions). Results are thresholded at p<0.05 voxelwise, with cluster-level FWE correction at p<0.05, masked to the relevant network mask. Color bars represent z-scores, and result maps are superimposed on the MNI brain template. Abbreviations: DMN = default mode network; CDR^®^ plus NACC FTLD-SB = CDR Dementia Staging Instrument plus Behavior and Language domains from the National Alzheimer’s Coordinating Center (NACC) Frontotemporal Lobar Degeneration Module, sum of boxes; GMV = gray matter volumes; MPN = medial pulvinar network; NfL = plasma neurofilament light chain; proC9 = prodromal *C9orf72* expansion carriers; SMN = sensorimotor network; SN = salience network; SxC9 = symptomatic *C9orf72* expansion carriers.

For SxC9, greater symptom severity at baseline (higher CDR^®^ plus NACC FTLD sum of boxes scores) correlated with longitudinal connectivity decreases (Figure 5B). These included longitudinal decreases in the SMN in the right pre- and post-central gyrus and bilateral midcingulate cortex, in the DMN in the right middle temporal gyrus, and in the MPN in the left pre- and post-central gyrus.

## 4 Discussion

Leveraging a large longitudinal cohort of *C9orf72* expansion carriers, we characterized functional and structural changes across asymptomatic, prodromal, and symptomatic stages. Despite no detectable longitudinal GM loss in any carrier group, we discovered longitudinal connectivity changes as early as the asymptomatic stage, even within a mean timeframe of 2.22 years. Importantly, the prodromal stage heralds a critical transition from the asymptomatic to symptomatic phase, yet previous studies have not characterized its functional connectivity profiles. Here, we identified in proC9 reduced baseline connectivity in regions of the SMN and enhancements in the DMN and MPN. Although proC9 did not exhibit group-level longitudinal connectivity changes relative to controls, greater baseline symptom severity correlated with longitudinal SN and SMN connectivity changes. Additionally, we identified associations between longitudinal connectivity changes and measures of neurodegeneration and clinical decline unique to each clinical stage. Higher baseline NfL was associated with longitudinal connectivity changes in the SMN and MPN in aSxC9, and GM decline and DMN connectivity increases in proC9/SxC9. Greater baseline symptom severity was associated with longitudinal connectivity changes in distinct ICNs for proC9 and for SxC9. In sum, our findings reveal stage-specific changes in longitudinal functional connectivity despite the lack of detectable gray matter atrophy declines, offering novel insights into network-level changes underlying *C9orf72*-related disease progression.

### Despite lacking gray matter decline, asymptomatic carriers demonstrate longitudinal connectivity changes in *C9orf72*-relevant networks which show associations with NfL

At baseline, aSxC9 showed SN, SMN and MPN connectivity disruptions, consistent with previous findings in asymptomatic carriers [2]. These networks have been implicated in cross-sectional studies of *C9orf72*-bvFTD (with or without MND), which shows SN and SMN alterations [20,52–54]. The medial pulvinar, which participates in social cognition and visual salience mechanisms [22], is distinctly atrophied in *C9orf72*-FTD [55] and *C9orf72*-ALS [14,28,56], and shows abnormally low GMV in aSxC9 [2]. For the DMN, we found no differences in aSxC9 compared to controls, contrasting with our previous study [2]. Previous cross-sectional analyses identified DMN hypoconnectivity during the asymptomatic phase [2], hyperconnectivity in early-stage or slowly progressive symptomatic carriers, and no differences in *C9orf72*-bvFTD [8]. These findings suggest that DMN connectivity is dynamic across clinical stages. One possibility is that our aSxC9 cohort (which included more than double the participants compared to previous studies) may have included more participants closer to symptom onset and thus, DMN “normoconnectivity” in our cohort may represent an averaging of the transition from DMN hypo- to hyper-connectivity across individuals.

Longitudinally, SN and MPN connectivity decreases were observed, which aligns with a study in which asymptomatic carriers had regions of longitudinal SN and thalamic network connectivity decline [9]. In the DMN, we identified regions of connectivity increases, in line with previous work showing age-related DMN increases in asymptomatic carriers [11].

Hyperconnectivity has been noted as an early response in various neurological diseases, including neurodegeneration, followed by diminishing connectivity as networks degrade in parallel with advancing atrophy [57–59]. Thus, one possibility is that the DMN connectivity increases represent a response to network disintegration of the remaining networks.

For the SMN, we did not identify longitudinal changes. However, one study reported both connectivity decreases and increases in a motor network [9], and another study found SMN increases with age [11]. These discrepancies in SMN connectivity across studies may reflect differences in study cohorts undergoing dynamic changes occurring during the asymptomatic phase. Moreover, these longitudinal studies consist of group-level findings such that cohort differences—particularly in the timing of future symptom onset—may contribute to the observed variability.

Next, we examined the extent to which longitudinal connectivity changes in aSxC9 are associated with a marker of neurodegeneration, and found that higher baseline NfL was associated with longitudinal SMN and MPN changes. Higher NfL was associated with longitudinal SMN decline in the right precentral gyrus. Interestingly, each of the three clinical stages featured regions of SMN hypoconnectivity at baseline. Taken together, these findings suggest that the longitudinal SMN decline seen in aSxC9 coincides with early neurodegeneration. For the MPN, higher NfL correlated with longitudinal increases within the right anterior thalamus and declines in bilateral temporoparietal regions. These divergent MPN changes appear to accompany early neurodegeneration, perhaps reflecting a combination of region-specific vulnerability and compensatory reorganization. Altogether, we propose that these NfL-associated SMN and MPN connectivity changes indicate neural responses to emerging neurodegeneration during the asymptomatic stage.

### In prodromal carriers, SN and SMN changes point toward risk of further decline

For *C9orf72*, there remain substantial barriers to identify carriers at greatest risk for symptomatic conversion. The age of symptom onset and rate of progression are highly variable, even within families [7,60]. *C9orf72-*bvFTD presents with insidious behavioral changes [61], which are challenging to quantify. Nevertheless, identifying which prodromal carriers are at greatest risk for symptomatic progression is crucial for treatment initiation considerations.

To our knowledge, this study is the first to assess network connectivity profiles during the prodromal stage. Cross-sectional analyses revealed connectivity alterations in the SMN, DMN and MPN. As in aSxC9, proC9 showed SMN disruptions at baseline. DMN hyperconnectivity was observed in proC9, which may reflect a continuation of the longitudinal DMN increases budding during the asymptomatic stage. ProC9 exhibited MPN hyperconnectivity in bilateral precuneus/posterior cingulate cortex, contrasting with our findings in aSxC9 which showed regions of MPN hypoconnectivity, including the precuneus, at baseline.

One possibility is that regions of DMN and MPN hyperconnectivity in proC9 represent a compensatory response to early neurodegeneration during this critical transition from the asymptomatic to symptomatic stage.

Although group-level longitudinal connectivity changes were not observed in proC9, we found that greater baseline symptom severity in this group correlated with longitudinal SN declines in bilateral medial frontal cortex and SMN increases in bilateral precentral gyrus. For the SN, declines appear driven by the proC9 individuals with greater symptom severity, which points toward declining SN connectivity as a sign of greater risk for clinical progression.

Similarly, regions of increasing SMN connectivity in proC9 may portend a greater risk of progression. Our data suggest a non-linear SMN trajectory across disease stages with reductions (vs. HC) found in all three stages, with longitudinally increasing SMN connectivity in SxC9, perhaps emerging as early as the proC9 stage in a subset of those with the greatest symptom severity. Once sufficient long-term longitudinal data have been collected from the prodromal to symptomatic stage, future studies will elucidate these trajectories.

### Symptomatic carriers predominantly have baseline hypoconnectivity with longitudinal connectivity network increases

Consistent with our prior study, SxC9 had baseline hypoconnectivity within key regions of the SN and SMN [8]. Unexpectedly, we found longitudinal SN and SMN connectivity increases in this group. For the SMN, increases in perirolandic and supplementary motor areas arose within regions that had disrupted connectivity at baseline. These findings contrast with a study showing that carriers predominantly with ALS or FTD-ALS lacked longitudinal connectivity changes [11]. The predominance of bvFTD phenotype and longer follow-up in our SxC9 cohort may explain this discrepancy.

In SxC9, greater baseline symptom severity was associated with longitudinal SMN declines in bilateral supplementary motor area, regions which also showed disrupted connectivity at baseline. This result points toward the notion that individuals at more advanced clinical stages show progressive SMN disruption. SxC9 exhibited longitudinal SMN increases in perirolandic regions, albeit regions that minimally overlapped with those showing SMN declines associated with greater baseline symptom severity. One potential explanation for this apparent paradox is that the group-level SMN increases may be driven by less impaired individuals, whereas those with more severe baseline symptoms are driven toward future SMN breakdown.

For the DMN, SxC9 showed regions of both hyperconnectivity and hypoconnectivity at baseline. Divergent DMN alterations have been described in bvFTD [52], the predominant clinical phenotype of the SxC9 cohort. Though a previous group-level analysis of *C9orf72*-bvFTD identified no DMN alterations, single-subject analyses of early-stage *C9orf72*-bvFTD revealed DMN hyperconnectivity [8], suggesting that individual heterogeneity could account for group-level discrepancies across studies. Notably, baseline DMN hyperconnectivity in SxC9 largely overlapped with hyperconnectivity in proC9 in the precuneus/posterior cingulate cortex. These regions represent key DMN hubs and were distinct from the region showing DMN declines associated with greater baseline symptom severity (right middle temporal gyrus). We propose that the baseline parietal DMN hyperconnectivity may reflect a reactionary, or potentially compensatory, mechanism related to neurodegeneration which becomes unsustainable with disease progression, as suggested by the absence of longitudinal DMN increases.

The medial pulvinar of the thalamus represents a key region of degeneration in *C9orf72*-bvFTD and -ALS [14,28,55,56]. Reduced connectivity to this region in *C9orf72-*bvFTD and - ALS [8,15], and asymptomatic carriers has been reported [2]. In contrast, our SxC9 did not show altered baseline and longitudinal MPN connectivity. Despite the lack of group-level differences, greater baseline symptom severity correlated with longitudinal MPN decline, suggesting that MPN changes are associated with symptom progression.

In a combined proC9 and SxC9 group, higher baseline NfL correlated with longitudinal precentral gyrus atrophy (a key hub of the SMN) and midcingulate cortex, in line with a previous study [62]. Moreover, higher baseline NfL was associated with longitudinal DMN increase in the angular gyrus, a potential reactionary response.

Longitudinal GM declines were not detected in any group. Similarly, previous studies report no detectable GM decline in asymptomatic carriers [11,25,29]. Given the slow atrophy rate reported even in symptomatic carriers [63,64], the relatively short mean follow-up time likely limited the ability to detect atrophy changes. In contrast, connectivity changes were captured during the study’s time frame, highlighting its potential for tracking dynamic changes longitudinally during each clinical stage.

### Conclusions and limitations

To our knowledge, this is the largest study of longitudinal functional connectivity changes in *C9orf72* expansion carriers, which enabled us to examine changes unique to the asymptomatic, prodromal and symptomatic stages. We identified stage-specific associations between ICN changes and clinical measures of neurodegeneration and symptom severity, even in the absence of detectable longitudinal GM loss. These results highlight tf-fMRI as a promising method for detecting change across *C9orf72* clinical stages, despite the relatively limited follow up time in the present study. Limitations of the present study include limited NfL data and limited participants with *C9orf72*-associated ALS, which precluded subgroup comparisons with *C9orf72-*bvFTD. Future studies with larger cohorts and longer follow up will be needed to establish connectivity trajectories throughout the lifespan and among distinct clinical phenotypes.

## Data Availability

This study's data are available upon reasonable request.

## Acknowledgements

This work was supported by the following agencies: NIH (S.E.L.: R01 AG058233 [NIA], R01AG071756 [NIA]; M.L.G.-T.: K24DC015544 [NIDCD], RF1NS050915 [NINDS], R01AG075775 [NIA]; J.S.Y.: R01AG062588, R01AG057234, P30AG062422, and U19AG079774 [NIA], U54NS123985 [NINDS]; V.E.S.: R01AG052496 [NIA]; B.F.B.: P30 AG062677 [NIA]; A.L.B.: R01AG078457, R01AG073482, R56AG075744, R01AG038791, RF1AG077557, R01AG071756, U24AG057437 [NIA]; B.F.B, A.L.B., K.K., E.M.R.: U19AG063911 [NIA]; M.L.M., M.L.G.-T., J.S.Y., V.E.S., A.L.B., E.M.R., H.J.R., W.W.S., T.M.F., S.E.L.: P01AG019724 [NIA]), the Tau Consortium (S.E.L., J.S.Y., B.F.B., A.L.B.), Päivikki and Sakari Sohlberg Foundation (S.H.), AlzOut and John Douglas French Foundation (J.C.R.), the Bluefield Project to Cure Frontotemporal Dementia (S.E.L., J.S.Y., A.L.B.), and the Alzheimer’s Association (A.L.B.). J.S.Y. also receives funding from the Global Brain Health Institute, Genentech, the French Foundation, and the Mary Oakley Foundation. C.U.O. receives the Jane Tanger Black Fund for Young-Onset Dementias, and the Nancy and Robert Hall Fund for Brain Research. A.L.B. also receives funding from the GHR Foundation, Association for Frontotemporal Degeneration, Gates Ventures, and Alzheimer’s Drug Discovery Foundation. K.K. was also funded by the Kathrine B. Andersen Endowed Professorship.

Data collection and dissemination of the data presented in this manuscript was supported by the ALLFTD Consortium (U19: AG063911, funded by the National Institute on Aging and the National Institute of Neurological Diseases and Stroke) and the former ARTFL & LEFFTDS Consortia (ARTFL: U54 NS092089, funded by the National Institute of Neurological Diseases and Stroke and National Center for Advancing Translational Sciences; LEFFTDS: U01 AG045390, funded by the National Institute on Aging and the National Institute of Neurological Diseases and Stroke). The manuscript has been reviewed by the ALLFTD Executive Committee for scientific content. The authors acknowledge the invaluable contributions of the study participants and families as well as the assistance of the support staff at each of the participating sites. Samples from the NCRAD, which receives government support under a cooperative agreement grant (U24AG021886) awarded by the NIA, were used in this study.

## Conflict of interest statement

C.U.O. has grant funds from Alector and Denali, and has served as consultant to Alector, Otsuka, Eli Lilly, Sanofi, Reata, and Neuvivo. J.C.R. reported serving as a site PI for clinical trials sponsored by Eli Lilly, Eisai and Amylyx; receiving consulting fees from Ferrer International, Adept Field Solutions and REACH. J.S.Y. serves on the scientific advisory board for the Epstein Family Alzheimer’s Research Collaboration and the Charleston Conference on Alzheimer’s Disease and is the editor-in-chief of npj Dementia. B.F.B. has received honoraria for SAB activities for the Tau Consortium - funded by the Rainwater Charitable Foundation; and institutional research grant support for clinical trials from Alector, Transposon, Cognition Therapeutics, EIP Pharma/Cervomed. A.L.B. has served as a paid consultant to Alector, Alexion, Arrowhead, Arvinas, Eli Lilly, Janssen, Merck, Neurocrine, Novartis, Oligomerix, Ono, Oscotec, Switch and Transposon. He is a scientific cofounder of Neurovanda. His institution received research support from Biogen and Eisai for serving as a site investigator for clinical trials, as well as from Regeneron. K.K. consults for Biogen Inc., Eisai Inc., and BioArctic Inc. with no personal compensation; She received research material support form Eli Lilly. H.J.R. has received grant funding from the NIH and the CA department of public health, and is a consultant for Eli Lilly and Alector. B.L.M. serves on the Scientific Advisory Board of the Bluefield Project to Cure FTD; the John Douglas French Alzheimer’s Foundation; Fundación Centro de Investigación Enfermedades Neurológicas, Madrid, Spain; Genworth, Inc.; the Kissick Family Foundation; the Larry L. Hillblom Foundation; and the Tau Consortium; serves as a scientific advisor for the Arizona Alzheimer’s Consortium and the Stanford University ADRC; receives royalties from Cambridge University Press, Elsevier, Inc., Guilford Publications, Inc., Johns Hopkins Press, Oxford University Press, and the Taylor & Francis Group; serves as editor for Neurocase and section editor for Frontiers in Neurology; and receives grants from the Bluefield Project to Cure FTD and NIH: P0544014, R01AG057234. L.Z., S.H., Y.J., M.L.M., D.L., J.L., M.L.G.-T., V.E.S., J.K., L.K.F., H.W.H., E.M.R., W.W.S., T.M.F., S.E.L. do not have any conflicts to disclose.

## Consent statement

This study was approved by institutional review boards, and was performed in accordance with the Declaration of Helsinki. All participants or their surrogates provided informed consent.

## Notes

### Author Declarations

Ethics committee/IRB of UC San Francisco gave ethical approval for this work. Ethics committee/IRB of Johns Hopkins University (Trial Innovation Network) gave ethical approval for this work.

