## Supplement for "Longitudinal functional network connectivity changes across the clinical stages of *C9orf72* hexanucleotide repeat expansion carriers"

**Supplementary Figure 1. Seed-based intrinsic connectivity networks of interest in *C9orf72* expansion carriers.** We investigated four networks of interest previously known to show cross-sectional alterations in symptomatic and asymptomatic *C9orf72* expansion carriers, including the SN, SMN, DMN and MPN. These network maps are applied as masks to constrain subsequent seed-based connectivity analyses. DMN = default mode network; MPN = medial pulvinar network; SMN = sensorimotor network; SN = salience network.

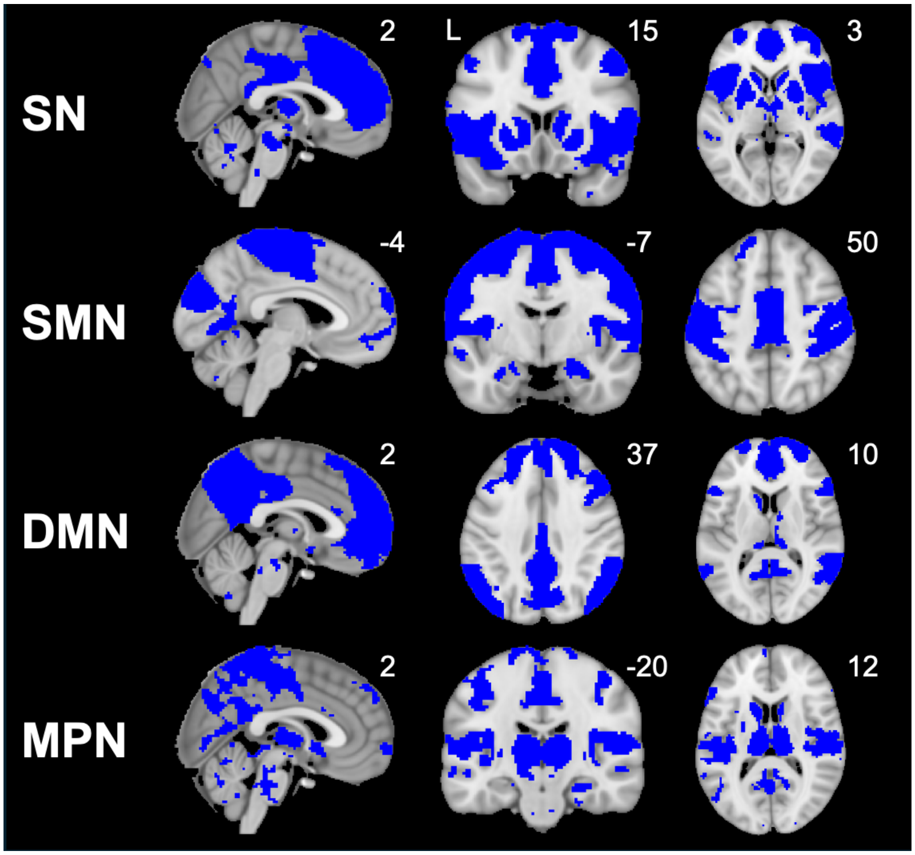

**Supplementary Table 1: Demographics and clinical measures for healthy controls used for ComBat harmonization and network masks**

| | **Combined study cohort (n=189)** | **Combat HC (n=83)** | **Seed mask HC (n=103)** |
|---|---|---|---|
| Age (years) | 51.7 (14.9) | 56.2 (16.4) | 55.4 (15.2) |
| Education (years) | 15.7 (2.4) | 16.0 (2.5) | 16.7 (2.9) |
| Sex (F:M) | 114:75 | 62:21 | 73:30 |
| Handedness (R/L/A) | 168/17/4 | 67/11/5 | 86/12/5 |
| CDR® plus NACC FTLD, global score, median [range] | 0 [0-3] | 0 [0] | 0 [0] |
| CDR® plus NACC FTLD, sum of boxes | 1.5 (3.5) | 0 [0] | 0 [0] |
| MMSE, total score | 28.3 (2.3) | 29.3 (0.9) | 29.4 (0.8) |
| MoCA, total score | 26.3 (4.2) | 27.1 (1.9) | 27.2 (1.9) |
| NfL (pg/mL) | 14.7 (24.1) | 7.0 (3.9) | 7.0 (3.8) |

Supplementary Table 1 presents group differences in the mean (SD) values (continuous variables) or proportions (categorical variables), unless otherwise specified. Original NfL concentration values are shown, while statistical analyses are based on a linear regression models using log-transformed NfL concentrations, which regress baseline age and sex as nuisance covariates. Abbreviations: CDR® plus NACC FTLD = CDR Dementia Staging Instrument plus Behavior and Language domains from the National Alzheimer's Coordinating Center (NACC) Frontotemporal Lobar Degeneration Module; F/M = female/male; HC = healthy controls; MMSE = Mini-Mental State Examination; MoCA = Montreal Cognitive Assessment; NfL = plasma neurofilament light chain; R/L/A = right/left/ambidextrous; SD = standard deviation.

**Supplementary Table 2. Peak coordinates of seed regions for ICNs of interest in *C9orf72* expansion carriers**

| Network | Seed region | MNI (mm) | | |
|---|---|---|---|---|
| | | *x* | *y* | *z* |
| SN | right anterior frontoinsula | 34 | 20 | -10 |
| SMN | right precentral gyrus | 28 | -16 | 66 |
| DMN | right angular gyrus | 56 | -52 | 26 |
| MPN | left medial pulvinar thalamus | -9 | -28 | 3 |

Abbreviations: ICN = intrinsic connectivity network; DMN = default mode network; MPN = medial pulvinar thalamus network; SMN = sensorimotor network; SN = salience network.
